# Using Paired Agent-Based Simulations To Test Strategies For Limiting The Effects Of Epidemics

**DOI:** 10.1101/19014043

**Authors:** Margaret Armstrong, Flávio Codeço Coelho

## Abstract

Agent-based simulations are widely used nowadays in public health research for comparing different strategies for mitigating epidemics and for planning appropriate responses in the aftermath of crises in large urban areas because they can capture fine scale heterogeneities that may have important non-linear effects on the results. Given the costs of implementing strategies, decision-makers have to be convinced that the proposed treatment/procedure leads to a statistically significant improvement.

This paper presents an innovative method for constructing paired agent-based simulations where exactly the same set of random effects is applied to simulations with and without the treatment/procedure. Statistical Analysis of Variance distinguishes the sum of squares between groups (BSS) from the sum of squares within groups (WSS). Our aim was to filter out the within sum of squares (WSS) leaving only the sum of squares between the control group and the treatment group (BSS). We propose to filter out the WSS by constructing paired simulations because as is well known, when paired t-tests can be used, they are much more powerful than ordinary t-tests. Pearson’s Chi-squared goodness of fit, the Kolmogorov-Smirnov statistic and the Kullback-Leibler Divergence are then used to test whether the effect is statistically significant. This procedure has been tested on a case-study on the propagation of the Zika epidemic in Rio de Janeiro in 2015.

**Author summary:** Agent-based simulations are emerging as a powerful tool in computational biology because they can capture fine scale heterogeneities that can have important effects on the propagation of epidemics. *In silico* experiments can be used to test different strategies for mitigating epidemics quickly and inexpensively. Given the inherent variability from one simulation to another, it is difficult to statistically prove their effectiveness. We have developed a powerful method rather like paired t-tests, for testing whether a given treatment is statistically better than the control. We do this by generating paired simulations with exactly the same random variables in the control simulation and the one with a treatment. Using the terminology of analysis of variance, we want to filter out the sum of squares within the group, leaving only the sum of squares between the control and the treatment. This procedure has been applied to a case-study to see whether enclosing and air-conditioning the transport hub in Rio de Janeiro would have slowed down the propagation of Zika.

## 1 Introduction

Over the past 20 years, several infectious diseases have reached epidemic proportions in different parts of the world: SARS and MERS in Asia, Ebola in Africa and Zika in Latin America. As the Zika virus causes microcephaly and serious neurological damage in unborn babies [1], and Guillain Barré syndrome in adults [2], it is important to understand how it propagates from one region to another and also within cities, so as to control it more effectively. The Zika virus is transmitted by mosquitoes of the Aedes genus^1^. As mosquitoes only fly short distances and usually remain near where they hatched [5], [6], [7], [8], human movement is a key component of vector-borne epidemiology [9]. In particular, transmission is influenced by social connections and the transport system [10], [11], [12], [13].

### 1.1 Case-study that motivated our research

An unusually large number of cases of microcephaly occurred in northeastern Brazil early in 2015. A few months later cases started occurring in the city of Rio de Janeiro. Figure 1 shows the total number of cases^2^ per “bairro” (suburb) from October 2015 to January 2016. These cases occurred in the northern part of the city along an east-west line and then towards the city center, apparently following the main transport routes. After 4 months none had occurred in the southern part of the city (called “Zona Sul”).

**Fig 1.**
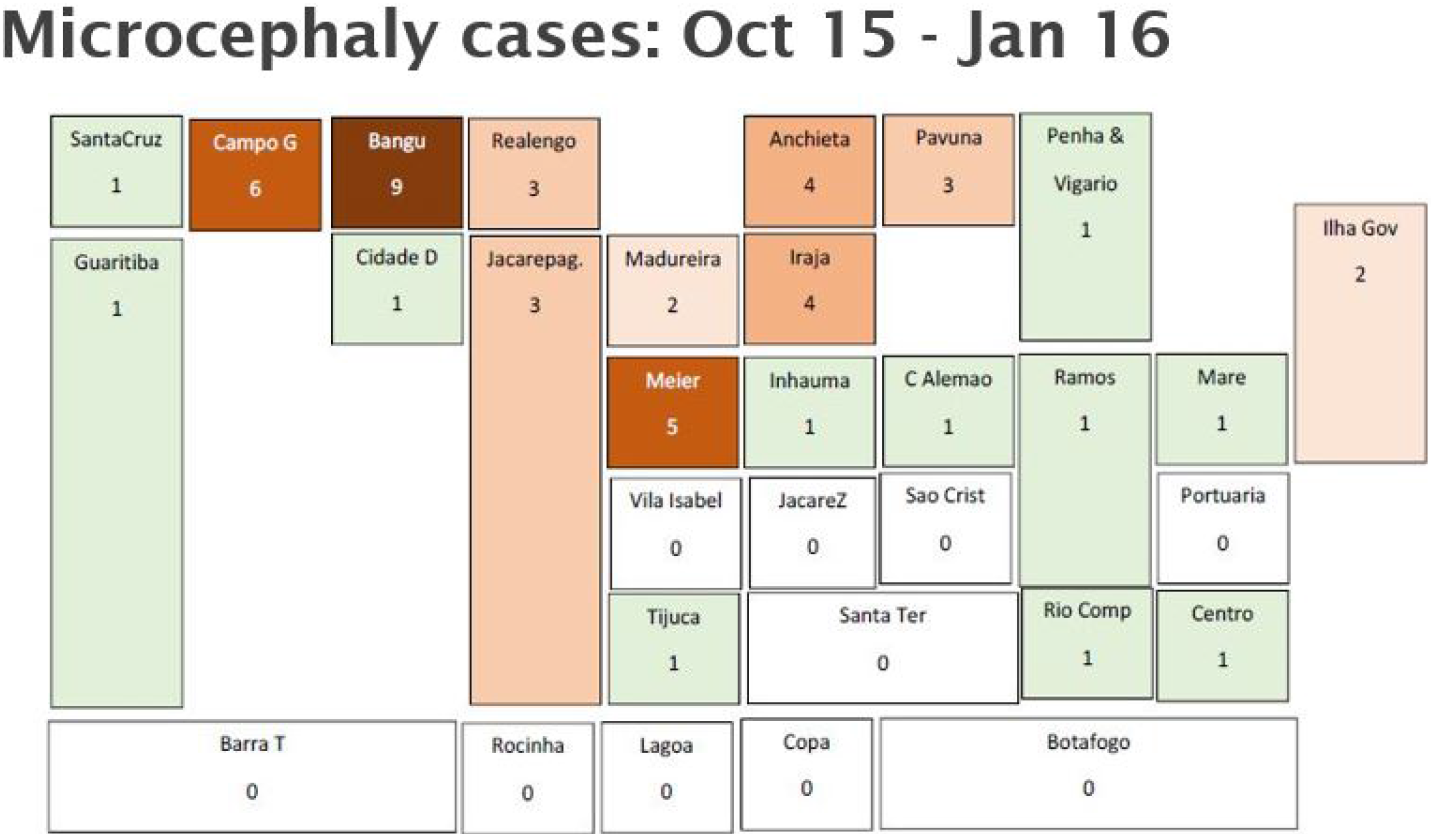
Number of cases of microcephaly from Oct 2015 to Jan 2016. Schematic representation of the layout of the 33 administrative regions (RAs) in Rio de Janeiro with the number of cases up to the end of January 2016. Cases are concentrated in the poorer northern suburbs especially Bangu and Campo Grande but there were none in the more affluent Zona Sul (Lagoa, Copacabana and Botafogo). As far as the favelas are concerned, there were no cases in Rocinha and Jacarezinho, and only 1 case in each of three other important favelas: Complexo da Maré, Complexe do Alemão and Cidade de Deus.

The objective of this research project was to model the propagation of the virus within this city with 6 million inhabitants, and in particular to understand what had delayed its arrival in Zona Sul because this might suggest ways to prevent the spread of vector-borne diseases. In the northern part of the city, the train and metro stations are above ground whereas in the southern part they are underground with forced ventilation in stations and tend to be mosquito-free. So our hypothesis is that people waiting at stations in the northern suburbs were often bitten by mosquitoes thereby facilitating the transmission of the virus, but not in the southern part. In that case, enclosing and air-conditioning stations in the northern part and the transport hub in the city center could significantly reduce the impact of mosquito-borne diseases: dengue fever, chikungunya, as well as Zika. Given the cost of enclosing and air-conditioning stations, the city authorities would want to be sure that it was cost-effective. So we needed a procedure for statistically testing this hypothesis.

### 1.2 Choosing the most appropriate model to use

The standard SEIR stochastic model [14] which assumes perfect mixing is clearly not appropriate in this case because the cases of Zika were preferentially located in certain areas, whereas our hypothesis is based on a detailed knowledge of mobility within the city. Recent work on the propagation of communicable/infectious diseases especially those transmitted by vectors, has emphasized the importance of taking account of the network structure of contacts [15] [16], of the dynamics of the mosquito populations [17], [18], [19], [20], [21], of the effect of local rainfall patterns [22], and of factors such as vaccination and immunity levels [23]. The papers [24], [25] concluded that agent-based simulations are better suited than standard approaches.

### 1.3 Comparing agent-based simulations without a treatment to those with the treatment

Our aim is to compare the number of cases of Zika, with and without treatment, as a function of time. Initially, we had run several hundred simulations under control conditions and independently the same number with the proposed treatment. As there was a huge variability within each set of simulations, it was not clear whether the difference between the mean of the control simulations and the mean of those with the treatment was due to the treatment or merely to the stochastic variability within each set of simulations. In statistics, Analysis of Variance distinguishes the sum of squares between groups (BSS) from the sum of squares within groups (WSS) [26]. Our aim was, if possible, to filter out the sum of squares within groups (WSS) leaving only the sum of squares between the control group and the treatment group (BSS). In classical statistics, it is well-known that when paired t-tests can be used, they are much more powerful than ordinary t-tests [27]. The innovative aspect of our work is the development of paired simulations in which exactly the same random effects are used in the simulations with and without the treatment. This procedure will be applied to agent-based simulations of Zika transmission within Rio de Janeiro, but it could be applied any other type of epidemic where social contacts are important.

In epidemiological models such as the SIR and SEIR models, the basic reproduction number *R*_0_ determines whether the epidemic should propagate (if *R*_0_ *>* 1) or die out naturally [14]. But in the presence of stochasticity the cutoff between the two regimes is not black and white, there is a grey zone where the epidemic could die out naturally [28]. Another innovation in this work is that the paired agent-based simulations provide an estimate of the probability of this occurring.

In determining whether a treatment is effective, the paired simulations can be divided into three classes:

1. Those simulations where the epidemic died out in both simulations. As these would not be seen in practice, they were eliminated from further study.
2. Those where the control simulation took off, but where the simulation with treatment was extinguished – clearly the desired result.
3. Those where both simulations took off. In almost all cases the one without treatment took off faster and affected a larger number of people

In our study on the propagation of the Zika epidemic in Rio de Janeiro, there were no cases where the simulation with treatment took off but the control one was extinguished.

The paper is structured as follows: Section 2 reviews agent-based simulations and their applications. In Section 3 we explain how to construct paired simulations. Section 4 reviews different statistics for testing whether the differences between the paired simulations are significant. In their study on the properties of different power divergence statistics, Cressie and Read [29] recommended using Pearson’s chi-squared goodness of fit if the distribution in the alternative hypothesis was peaked. They also noted that the Kullback-Leibler divergence is a constant multiple of the classic log likelihood ratio. A hypothesis test based on the Kullback-Leibler divergence using Hoeffding’s inequality was developed by Nowak^3^. In addition to these, we also propose to use the classic Kolmogorov-Smirnov statistic. Section 5 presents the case-study on Zika propagation originally presented in [30]. The results are discussed in section 6. Our conclusions and perspectives for future work are presented in Section 7.

## 2 Literature review on agent-based simulations

Agent-based simulations are widely used nowadays for modelling the propagation of infectious diseases, for comparing the different strategies for mitigating epidemics and for planning appropriate responses in the aftermath of crises in large urban areas, because they can capture fine scale heterogeneities that may have important non-linear effects on the results. In public health research, they have been used to evaluate different strategies for mitigating the severity of influenza-type pandemics [31–33], for modelling measles outbreaks [23], for modelling hospital-acquired infections [34], for determining the best way to allocate vaccines [35], [36] and for modelling the impact of sexual transmission of the Ebola virus [37]. Agent-based simulations have been used in research on malaria to model the infectious reservoir in humans [38] and to study the spatial and temporal heterogeneities of malaria incidence in a rainforest environment [39, 40]. Manore et al. [41] compared the transient and endemic behaviour of dengue and chikungunya in order to evaluate the risk of emergence of different virus-vector assemblages. Going further afield, these methods have been used to simulate infectious diseases in the immune system [42], biomolecular networks [43], myxobacterial development [44] and dosimetry in a virtual hepatic lobule [45]. Disaster preparedness policies and interventions in the event of a large human-initiated crisis such as a nuclear strike in an urban area have also been studied [46], [47].

The sensitivity of agent-based simulations to small changes in the parameter values was evaluated by using massive parallel computation and interactive data visualization [48]. Rizzi et al. [49, 50] have developed computationally efficient ways of simulating large epidemiological models by using a bit-string approach. Considerable work has been carried out to determine the best way to set up these models and how to scale them [33, 51–54].

### 2.1 How to test different strategies

In the literature, strategies are usually tested by comparing the values of a few statistics such as the attack rate and the reproduction number *R*_0_ computed after running a limited number of simulations [31, 32, 37]. For example, Dorratoltaj et al. [31], measured the attack rate, the reproduction number *R*_0_ and the economic costs for three strategies (no vaccine, static vaccination and dynamic vaccination) for three severity levels of the epidemic (catastrophic, strong and moderate) after running 25 simulations while Ferguson et al. [32] compared the attack rates and the reproduction numbers for different strategies. In the study on the Ebola virus, the average number of days during which the epidemic continued, was computed [37]. In many ways it would be more informative to study the effect of the epidemic over time.

## 3 Constructing paired simulations

This section is divided into two parts: first we describe the general procedure for generating agent-based simulations of the propagation of the Zika virus in Rio de Janeiro, then we explain how to construct paired agent-based simulations by extending the previous procedure. Those who are familiar with agent-based simulations and are primarily interested in constructing paired simulations can skip the first section.

### 3.1 Agent-based simulation of the propagation of the Zika virus

In order to simulate the propagation of the Zika virus within Rio de Janeiro, we constructed a simplified (“toy”) example of the transport system consisting of 5 suburbs (RAs):

- three in the northern suburbs along the above ground train lines (Bangu, Campo Grande and Meier) where many microcephaly cases had occurred,
- the city center with the main business area (Centro) and the transport hub
- and the southern suburbs (Zona Sul) where the metro is underground.

Following Barmak et al [12] and Barmak, Dorso and Otero [13] we divided each RA into two areas: the area around the station and the rest of the RA. This gave us 10 sub-zones each with its own mosquito and human population. According to the 2010 census, only 41,000 people live in the central area compared to about 400,000 in Bangu and in Meier, and about 550,000 in Campo Grande and in Zona Sul. So in our model there are no inhabitants in Centro. The populations in the other four RAs were divided into two groups: those that go to the city center to work each day and those who remain in their home area. The latter could go to work or school there, or could stay at home. People who stay in their residential area mix freely with each other, but not with those from other areas. In contrast, people who go to work in the business area, mix freely with other workers while doing business and while having lunch, and then again on the platform waiting to go home. Similarly, the mosquito populations are distinct and do not mix with mosquitoes from other areas.

#### 3.1.1 Interactions between mosquitoes and people

We divided the day into four periods: early morning when those who go to the city center wait on platforms and get bitten by mosquitoes), midday (when people are in contact either with those in their home area or with those in the city center), late afternoon when travellers wait on platforms at Centro and can be bitten by mosquitoes) and finally the evening (when everyone is at home and can be bitten by the mosquitoes there). Table 4 summarizes the interactions in the control case between the 10 mosquito populations and the 8 human populations over the course of each day. For example, the human populations P1, P3, P5 and P7 wait for a train home late in the afternoon and can get bitten by mosquitoes from the M9 population. In the evening the human populations P1 and P2 are at home in Bangu and can be bitten by mosquitoes from the population M2. Given the importance of the central transport hub in mixing people from different suburbs, the treatment that we consider is enclosing and air-conditioning it. To model this we eliminate the mosquito population at Centro.

**Table 1.**
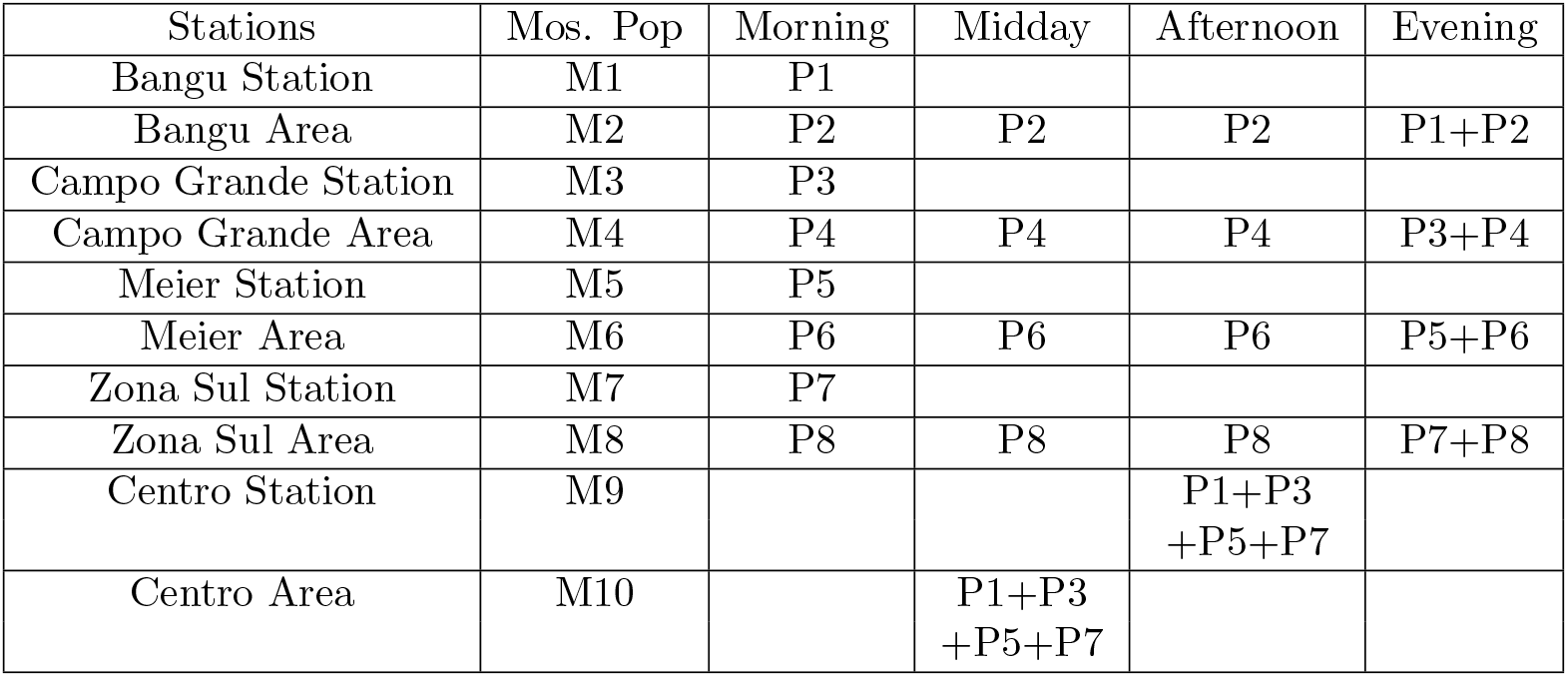
Location of human and mosquito populations in the control case. Human populations, P1, P3, P5 and P9, travel to work in the central business area during the day. Populations, P2, P4, P6 and P10, remain in their residential area all day. No people reside in the central area. The mosquito populations, M1 …M10 remain in their areas all day.

| Stations | Mos. Pop | Morning | Midday | Afternoon | Evening |
| --- | --- | --- | --- | --- | --- |
| Bangu Station | M1 | P1 |  |  |  |
| Bangu Area | M2 | P2 | P2 | P2 | P1+P2 |
| Campo Grande Station | M3 | P3 |  |  |  |
| Campo Grande Area | M4 | P4 | P4 | P4 | P3+P4 |
| Meier Station | M5 | P5 |  |  |  |
| Meier Area | M6 | P6 | P6 | P6 | P5+P6 |
| Zona Sul Station | M7 | P7 |  |  |  |
| Zona Sul Area | M8 | P8 | P8 | P8 | P7+P8 |
| Centro Station | M9 |  |  | P1+P3<br>+P5+P7 |  |
| Centro Area | M10 |  | P1+P3<br>+P5+P7 |  |  |

**Table 2.** Main parameters in model.

|  |  |
| --- | --- |
| Prob human to mosquito | Prob mosquito to human |
| 0.2 | 0.2 |
| Incubation period in humans | Incubation in mosquitoes |
| 5 days | 7 days |
| Daily mortality mosquitoes | Time before recovery in humans |
| 0.05 | 6 days |

**Table 3.**
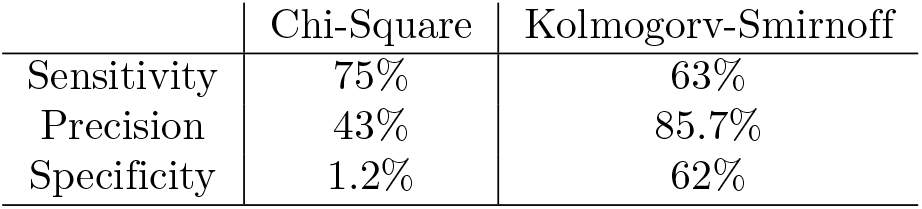
Sensitivity, Precision and Specificity for both tests.

**Table 4.** Human and mosquito populations at stations and in areas.

| Stations | Bangu | CGrande | Meier | Centro | Zona Sul |
| --- | --- | --- | --- | --- | --- |
| Mos. Pop | M1 | M3 | M5 | M7 | M9 |
| Morning | P1 | P3 | P5 |  | P9 |
| Midday |  |  |  |  |  |
| Afternoon |  |  |  | P1+P3+P5+P9 | Cell6 |
| Evening |  |  |  |  |  |
| Home areas | Bangu | CGrande | Meier | Centro | Zona Sul |
| Mos. Pop | M2 | M4 | M6 | M8 | M10 |
| Morning | P2 | P4 | P6 |  | P10 |
| Midday | P2 | P4 | P6 | P1+P3+P5+P9 | P10 |
| Afternoon | P2 | P4 | P6 |  | P10 |
| Evening | P1+P2 | P3+P4 | P5+P6 |  | P9+P10 |

#### 3.1.2 Updating at the end of each day

At the end of each day, after the four sets of interactions between the mosquitoes and people, the status of each individual agent is updated. For example, people who were exposed to the virus for the incubation period become infectious, and likewise for mosquitoes. Mosquitoes which “died” during the day are replaced by a new susceptible mosquito, etc.

Note these updating operations do not change the number of agents in each population; they can at most change their current state. This is important when setting up paired simulations because it makes it simpler to assign exactly the same random effects to the right individual agents. One does not have to worry about agents disappearing or new ones appearing.

The relationship between the four sets of interactions and the daily updating are shown in the flowchart in figure 2.

**Fig 2.**
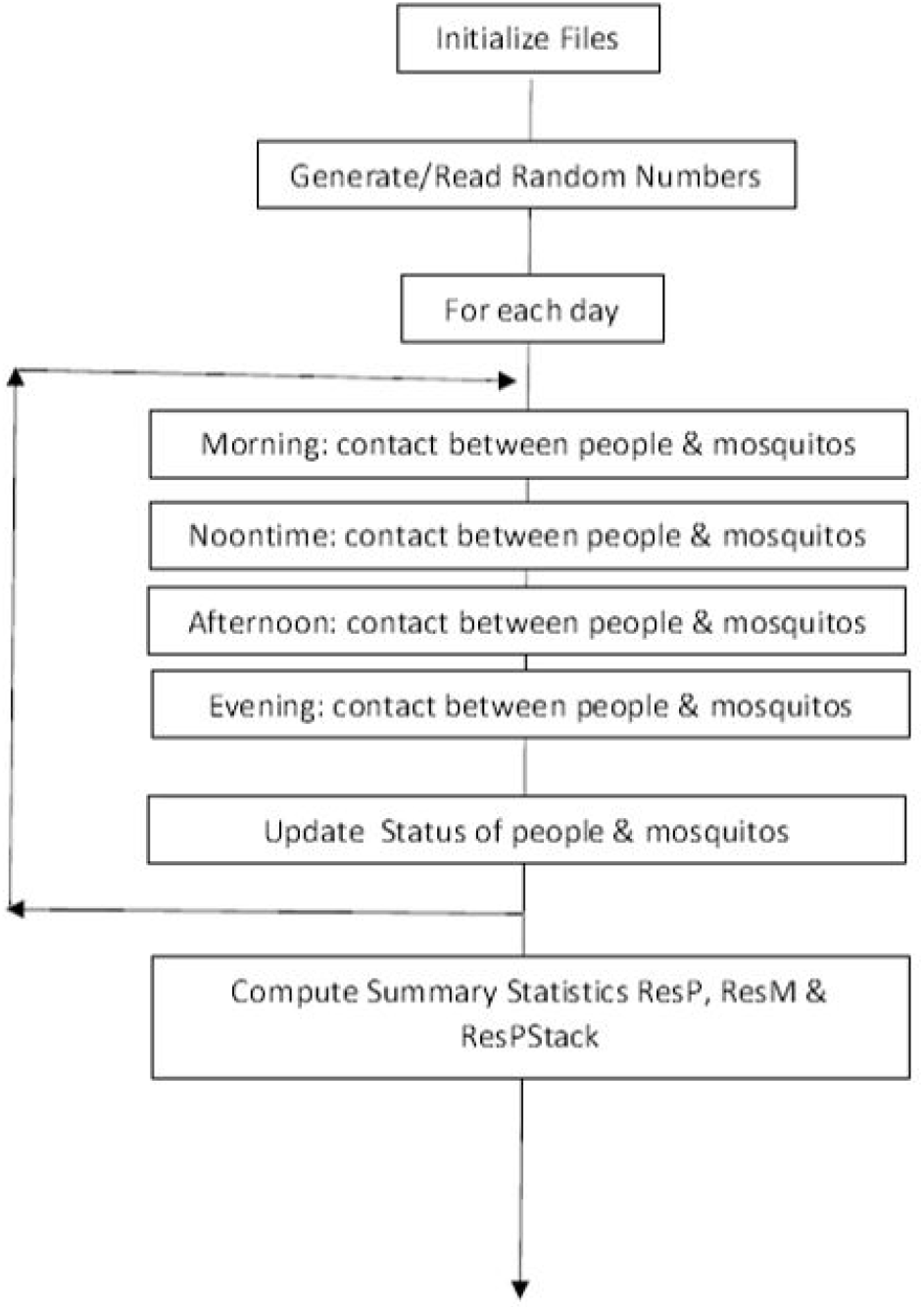
Schematic flowchart for generating agent-based simulations. This figure shows the key steps in the procedure for agent-based simulations of the propagation of the Zika virus in Rio de Janeiro.

#### 3.1.3 Parameters of the model

We assumed that, in the control case, each of the 10 mosquito populations consisted of NM=100 agents. Similarly there were a total of 1600 people, 200 in each of the 8 human populations. As the treatment that we wish to study consisted of enclosing Centro station and air-conditioning it to eliminate mosquitoes, the mosquitoes were removed from this station in the model of the treatment case.

As we consider an SEIR model for people and and SEI model for mosquitoes, the 6 key parameters are the incubation times for people and for mosquitoes (i.e. the time between being exposed to the virus and becoming infectious), the time for people to recover from the virus, the daily mortality rate for mosquitoes and the probability of the virus being transmitted to a susceptible person from an infectious mosquito, and similarly from a infectious person to a susceptible mosquito. As we only consider a period of 120 days, human mortality has been ignored.

The values of the main parameters in the model listed in Table 5 were taken from [55]. When a mosquito dies it is replaced by a new susceptible mosquito. We did not allow for vertical transmission of the virus between mosquitoes and their progeny. For simplicity, the incubation periods and the time to recovery were taken to be deterministic.

**Table 5.** Caption

| Prob human to mosquito | Prob mosquito to human | Daily mortality mosquitoes |
| --- | --- | --- |
| 0.2 | 0.2 | 0.05 |
| Incubation period in human | Time before recovery in humans | Incubation in mosquitoes |
| 5days | 6 days | 7 days |

### 3.2 Constructing paired agent-based simulations

The results of a simulation depend on the sequence of (pseudo-) random numbers that are used.The flowchart for each pair of agent-based simulation is shown on figure 3. Random numbers (or random variables) are required at each of the four time periods in each day (morning, noontime, etc), to decide which people are bitten by mosquitoes and whether they are infected.

**Fig 3.**
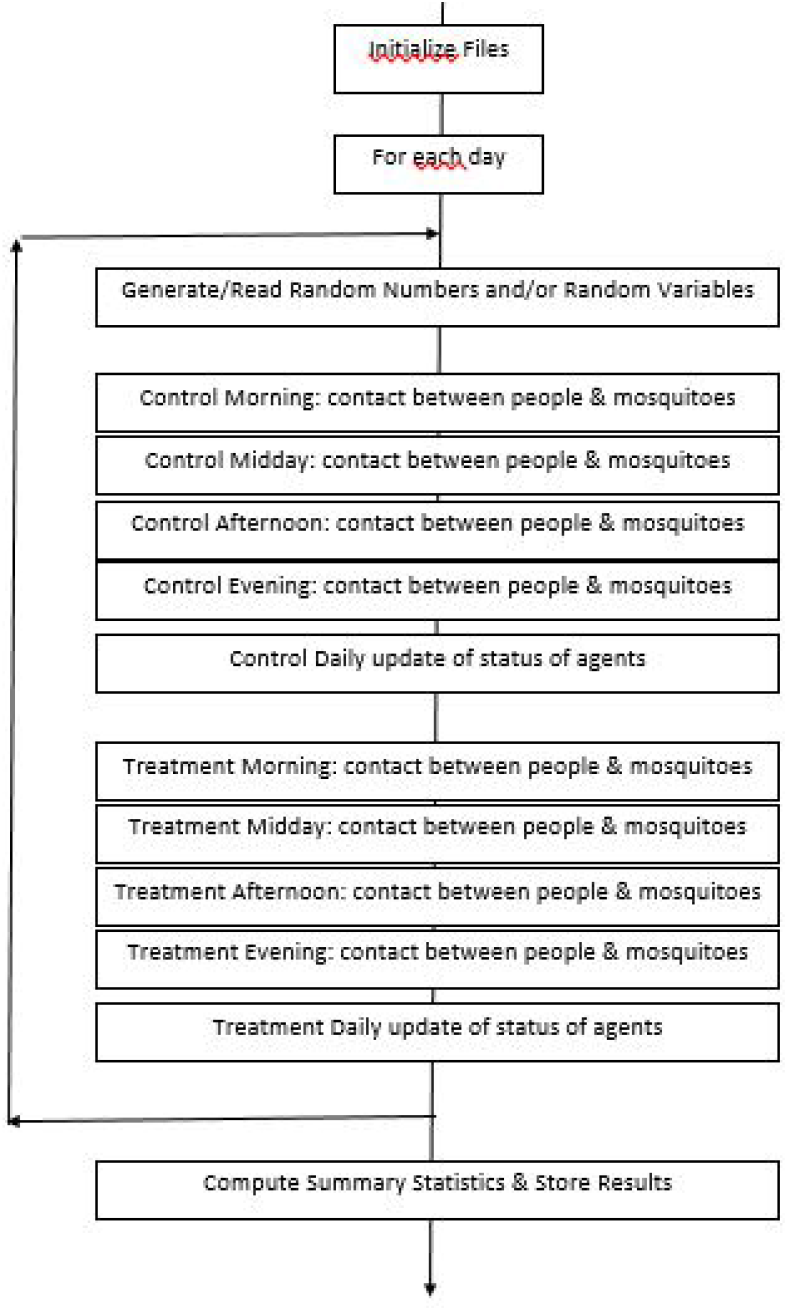
Schematic flowchart for generating paired agent-based simulations. This figure shows the key steps in the procedure for agent-based simulations of the propagation of the Zika virus in Rio de Janeiro. The full suite of programs used to generate the simulations on Github: https://github.com/fccoelho/PairedSimulations.

So the key to generating paired agent-based simulations is using exactly the same sequence of random numbers with and without the treatment. There are two ways of getting the same sequence: either by storing the sequence if it is not too long or alternatively by regenerating the exactly same sequence given the random seed, and the parameters of the algorithm. Until about 2005, linear congruential random number generators were the most commonly used generators [56]; nowadays the Mersenne twister [57] is more widely used. See [58] for a clear description of how the latter works.

## 4 Statistics for comparing the distributions

### 4.1 The Cressie-Read family of power divergences

Cressie & Read (1984) investigated a family of power divergence statistics, *I*(*λ*), for testing the fit of observed frequencies {*X*_*i*_; *i* = 1, …, *k*} to a theoretical distribution *π*_0_, or to expected frequencies *E*_*i*_ [29]. These are defined by

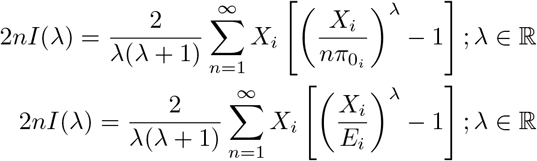

Pearson’s *χ*^2^ corresponds to the case *λ* = 1; the log likelihood ratio *G*^2^ is the case *λ* = 0; the Freeman-Tukey statistics *T* ^2^ corresponds to the case *λ* = −1*/*2; the modified log likelihood ratio *GM* ^2^ is the case *λ* = −1 while the Neyman modified *χ*^2^ corresponds to the case *λ* = −2. They noted that the log likelihood ratio is a constant multiple (twice) of the Kullback-Leibler divergence [59, 60]. After extensively testing these estimators, their recommendations were to use:

1. any *λ* ∈ [0, 1.5] when no knowledge about the alternative hypothesis is available
2. *λ* = 0 (i.e. the log likelihood ratio) if the alternative is dipped,
3. *λ* = 1 (i.e. Pearson’s *χ*^2^) if the alternative is peaked

As our alternative hypothesis is peaked, we use Pearson’s *χ*^2^ (*λ* = 1). As the log likelihood ratio *λ* = 0 is twice the Kullback-Leibler divergence and as Cressie and Read recommended using it when the alternative is dipped (not our case), we suspect that the Kullback-Leibler divergence will not be very powerful in our case.

### 4.2 Kullback-Leibler using Nowak’s hypothesis test

The Kullback-Leibler divergence is defined as

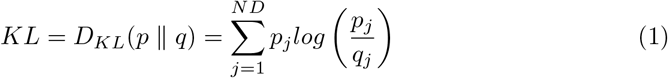

where *p* is the null hypothesis and *q* is the alternative hypothesis.

Using Hoeffding’s inequality [61], Nowak established bounds for the Type 1 and Type 2 errors, for two distributions, p and q, having the same support and such that

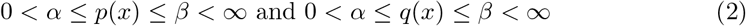

in terms of the two Kullback-Leibler divergences *D*^2^(*p* ‖ *q*) and *D*^2^(*q* ‖ *p*):

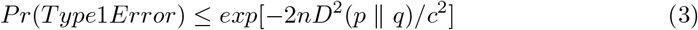

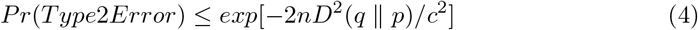

Here *c*^2^ = 2[*log*(*β*) −*log*(*α*)]

### 4.3 Kolmogorov-Smirnov hypothesis test

Hypothesis tests using the Cressie and Read’s family are all based on the multinomial distribution, and consequently they involve the expected and observed frequencies in predefined classes. Another way of looking at the problem is by comparing the cumulative number of cases over time. If these are expressed as a fraction of the total population, then classic tests such as the Kolmogorov-Smirnov test can also be used [62], [63], [64], [65].

The two-sample Kolmogorov-Smirnov statistic is

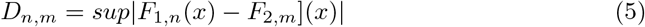

where *F*_1,*n*_(*x*) and *F*_2,*m*_ are the empirical distributions of the first and second samples. For large samples, the null hypothesis is rejected at level *α* if

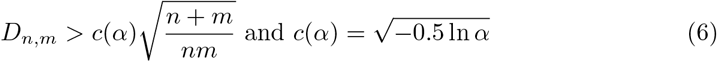

In particular, *c*(0.05) = 1.224.

## 5 Case study on Zika in Rio de Janeiro

To test these statistics we ran 200 sets of paired simulations of our model for the propagation of Zika in Rio de Janeiro. As this paper focuses on the methodology for testing whether the effect of the treatment is statistically significant, we will not present the case-study in detail here. It is summarized in Appendix 2 and is described in detail in [30].

In 36 out of the 200 sets of paired simulations, the epidemic died out in both the control case and the case with treatment. As these cases would never have been observed in the real world, they were removed from further study, leaving 164 pairs of simulations in the study. In 4 of these cases the treatment was quite effective; at most two cases occurred in addition to the two index cases. These four cases were treated separately. In Figure 4 the cumulative number of cases is shown in black in the control simulation and in red in the one with treatment.

**Fig 4.**
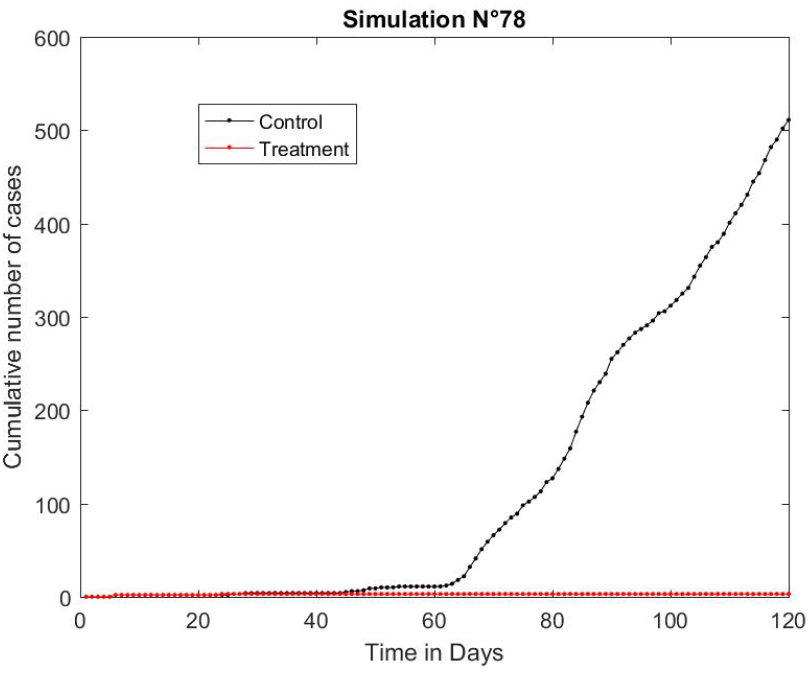
Paired simulations N°78. The cumulative number of cases up to 120 days are shown with the control case in black and the one with treatment in red. Even though the epidemic took off in the control case, it died out in the case with treatment, which is a very favourable result.

Figure 5 presents the cumulative number of cases for four typical pairs out of the remaining 160 paired simulations with control case shown in black and the one with treatment in red. Note how variable the results are. This variability makes it difficult to test whether the treatment is effective.

**Fig 5.**
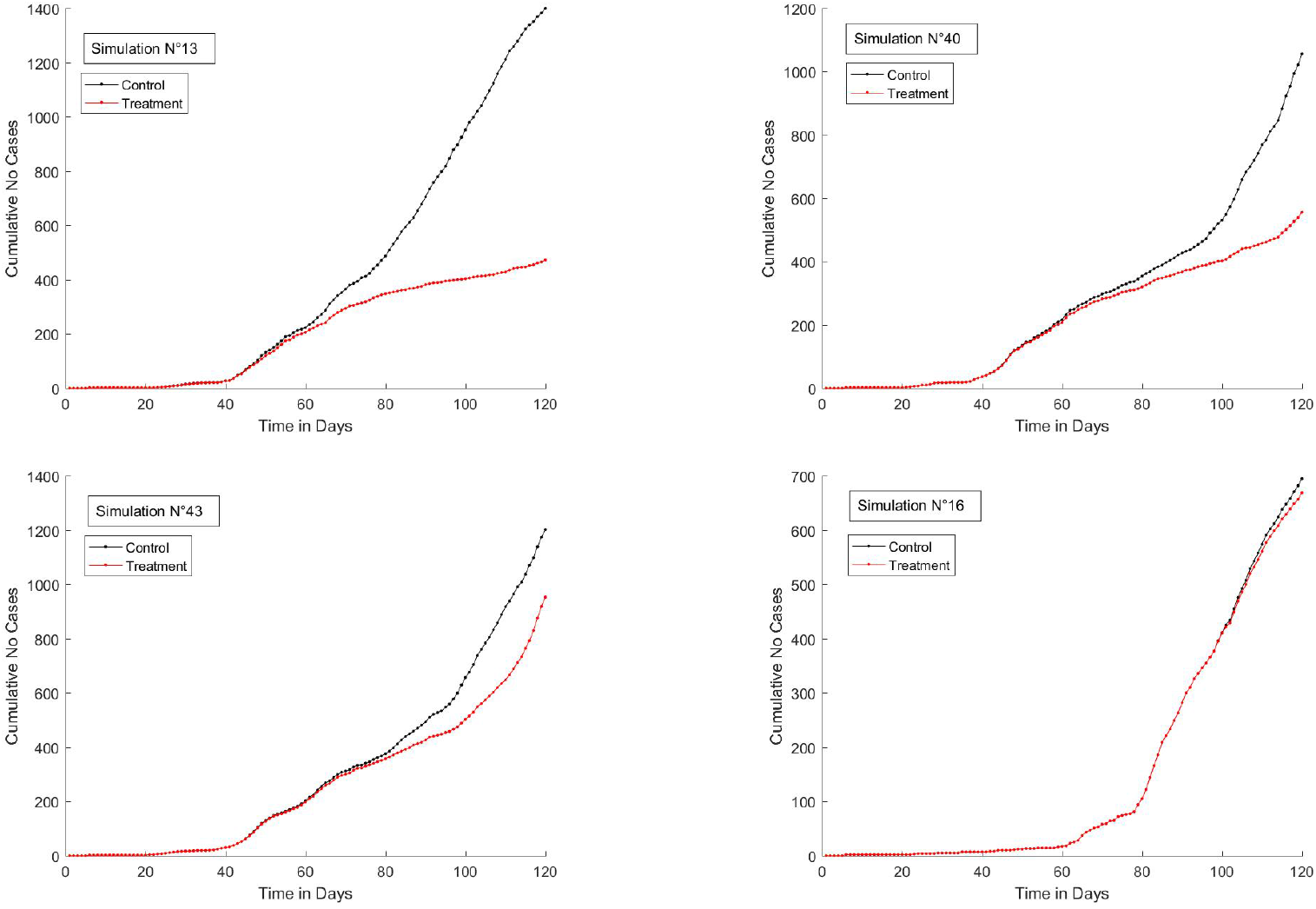
Four typical sets of paired simulations. Four typical sets of paired simulations showing the cumulative number of cases up to 120 days, with the control case in black and the one with treatment in red. Note how variable the simulations are.

### 5.1 Choosing the size of the classes for the tests

The simulations give the cumulative number of cases of Zika as a function of time up to 120 days (this corresponds to the summer period when mosquitoes reproduce in Rio de Janeiro). On many days there were no incremental cases in the control simulation.

Looking back at the formulas for the Chi-squared goodness of fit, we see that the expected number *E*_*i*_ has to be positive in all the classes. In addition for the Kullback-Leibler divergence, the number of observations *O*_*i*_ in each class must also be positive. So we defined the classes to have a minimum length of 6 days and if *E*_*i*_ or *O*_*i*_ is zero we increased the length until some new cases occurred. This led to 16 or 17 classes. Exactly the same classes are used for the two simulations in a pair (control and with treatment), but necessarily from one pair to another.

## 6 Results and discussion

### 6.1 Comparing the average cumulative curves

The standard way of testing whether a treatment is effective is by comparing the average number of cases for the the control case and the one with treatment. In Fig 6 the average of the control cases is shown in black, with the treatment in red. The values of the Chi2 goodness of fit and the KS statistics were 93.103 and 0.1734. As their critical values at the 95 % level of confidence are 26.30 and 0.1242, both are significant. The weak point about using averages is that it does take account of the variability of the individual cumulative curves around their respective averages. To illustrate this point, we superimposed the first 30 individual curves out of 160 in blue on top of the corresponding average curve, with the control case on the left and the treatment ones on the right (Fig 7).

**Fig 6.**
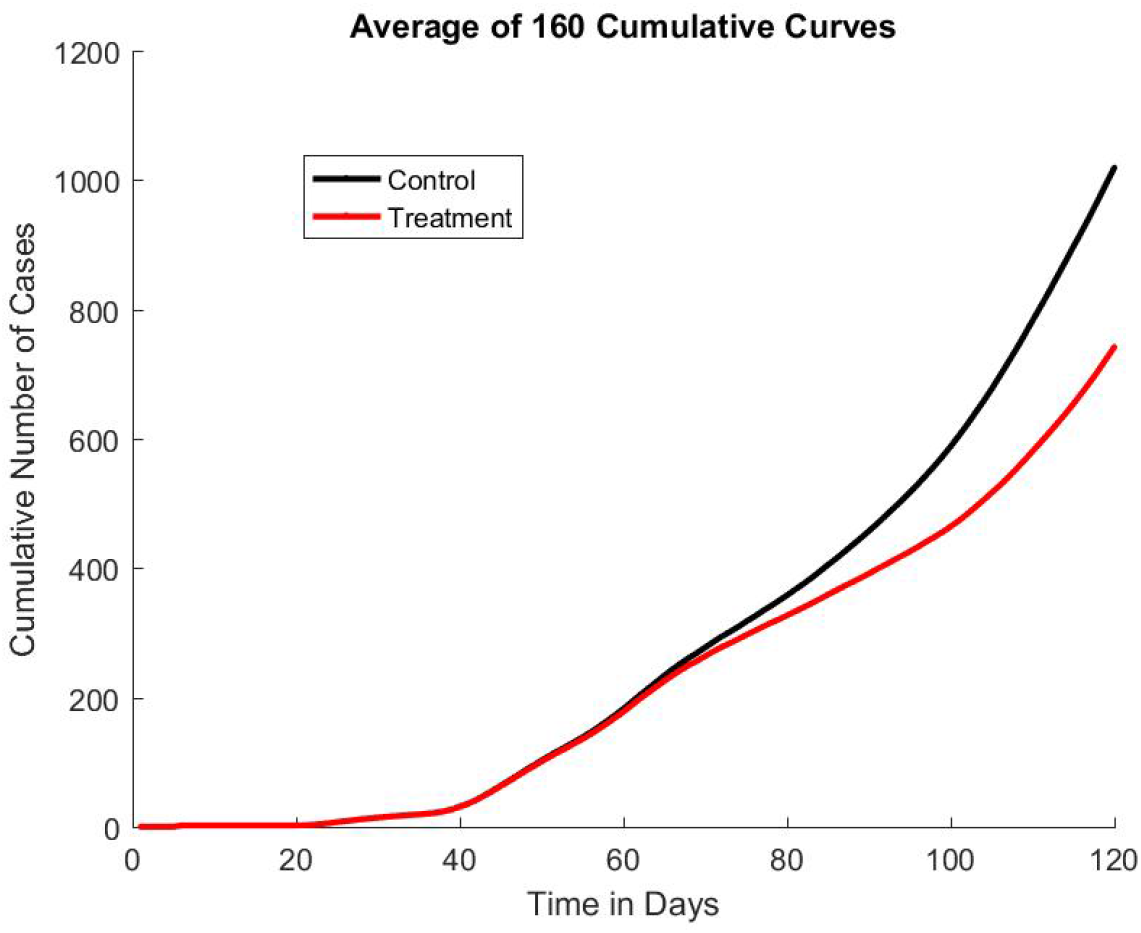
Average Number Cumulative Cases as a function of time. The control case is in black, and the one with treatment in red.

**Fig 7.**
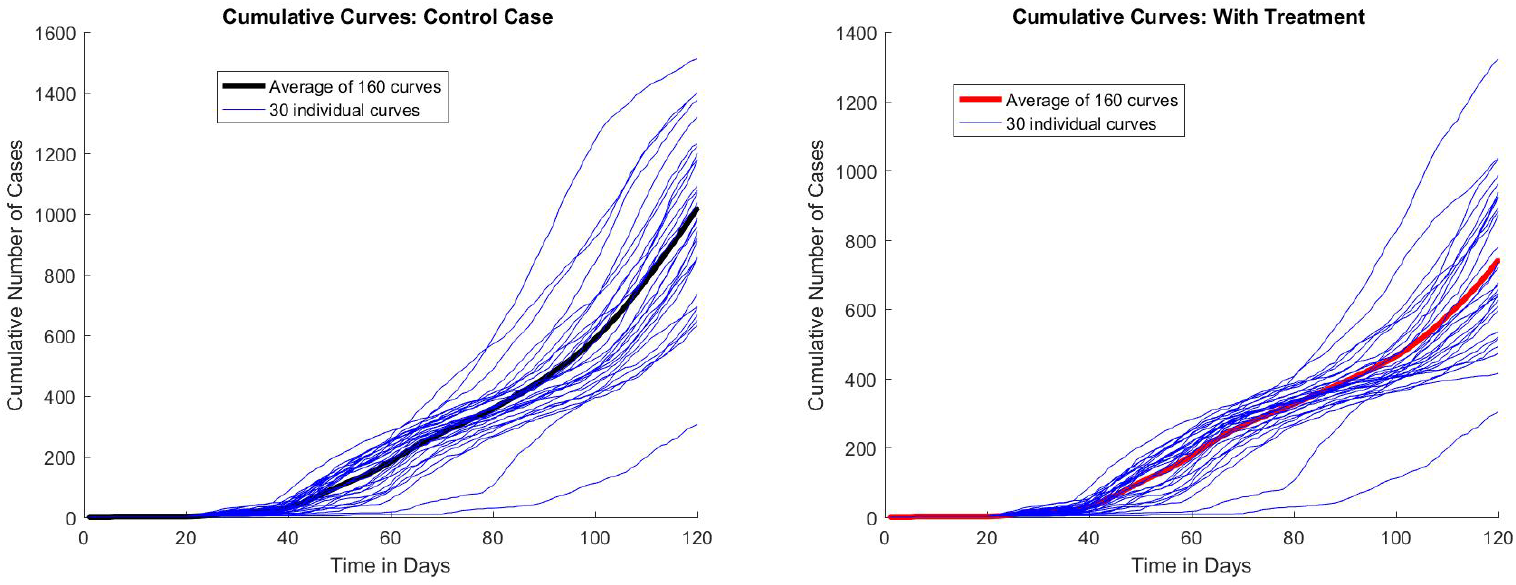
Average Number Cumulative Cases and 30 of the 160 simulations. The average of the 160 simulations is shown in bold, with the control case on the left and the one with treatment on the right. The first thirty individual simulations out of the 160 are in blue to show their variability.

### 6.2 Statistics for the paired simulations

Next we computed the statistics and determined how many sets of paired simulations exceeded the 95% confidence level. Nowak’s test for the Kullback-Leibler divergence performed very poorly. This is hardly surprising because it is a multiple of the log likelihood ratio and Cressie and Read had recommended using that only when the alternative distribution was dipped whereas it is peaked in our case. As was mentioned, the critical values for the KS statistic and the Chi2 goodness of fit at the 95 % level of confidence are 0.1242 and 26.30 respectively. In the KS case 86 out of the 160 sets of paired simulations exceeded this value, compared to 122 out of the 160 sets for the chi-squared test. In Fig 8 the values of the two statistics are presented. Empty circles correspond to pairs of simulations where neither statistic is significant; red circles, to cases where both are significant while the yellow circles refer to cases where the chi-squared goodness of fit was significant but the Kolmogorov-Smirnov was not.

**Fig 8.**
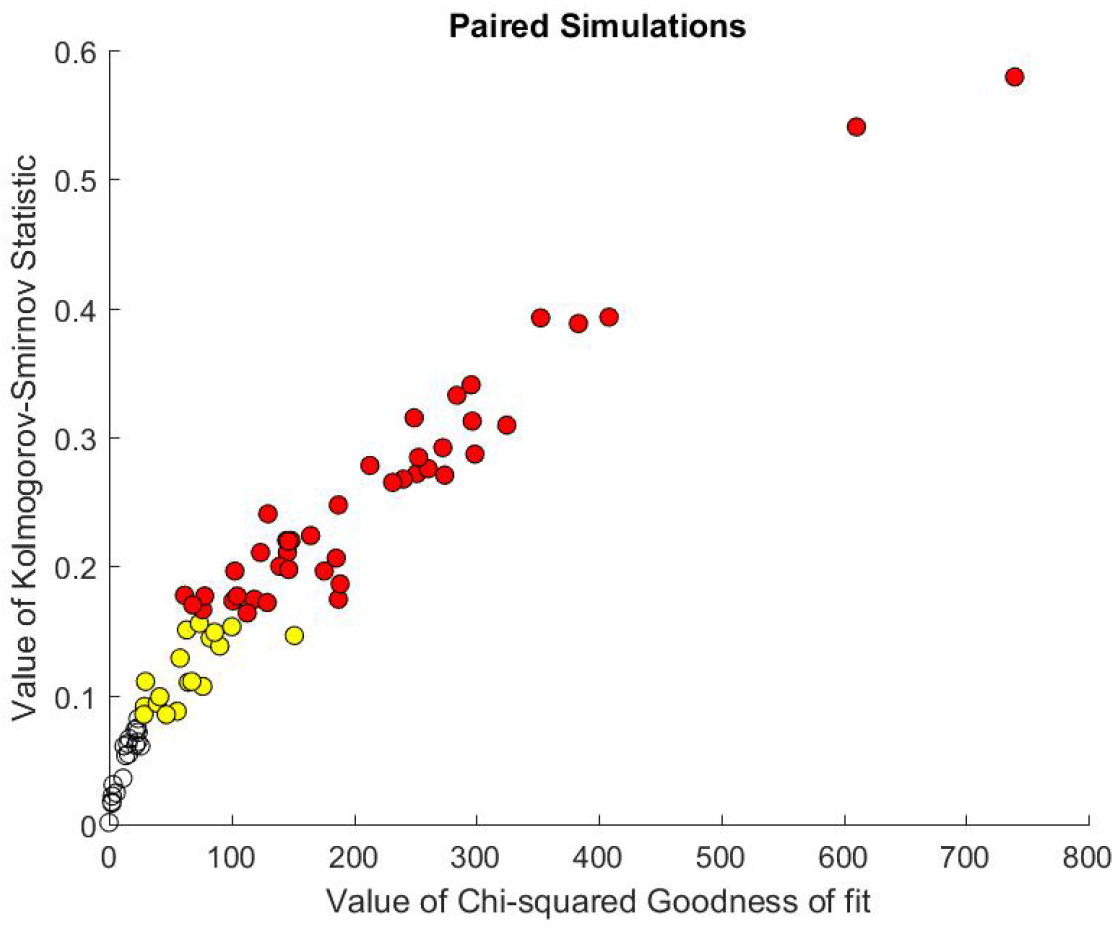
Kolmogorov-Smirnov statistic vs the chi-squared goodness of fit. The empty circles correspond to pairs of simulations where neither the chi-squared goodness of fit (Chi2) nor the Kolmogorov-Smirnov statistic (KS) are significant; the red circles, to cases where both are significant and the circles coloured yellow refer to cases where the chi-squared goodness of fit was significant but the Kolmogorov-Smirnov was not.

The results presented above indicate that the chi-square test is more sensitive than the KS. For a more complete assessment of the performance of both tests, we calculated the precision (or positive predictive value) and the specificity(or true negative rate) for both tests [66]. To calculate these metrics, we need to establish a few basic quantities for each test, namely the number of true positives(*tp*), i.e. the pairs (control,treatment) marked as different by the test; the number of true negatives (*tn*), i.e. the pairs (control,control) marked as not different by the test and the number of false positives(*fp*), i.e. pairs (control, control) marked as different by the test. We used 160 pairs of each kind (positives and negative) in these calculations.

Sensitivity is the fraction of (control, treatment) pairs correctly identified as different, 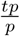. Precision, 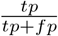, corresponds the fraction of pairs the test classifies as different (positive) which are really different. The specificity, 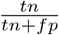, measures the proportion of actual negatives (control,control) that are correctly identified as such, in this case how often the simulations are actually the same when the test says so. The results from these calculations are presented in Table 3. From these metrics we see that the KS test although less sensitive, is much more precise and specific.

### 6.3 If the simulations had not been paired

In this section we consider what would have happened if the simulations had not been paired. In that case we could only have compared a control simulation with an arbitrary simulation with treatment. As we have a total of 160 of each type of simulation, there are a total of 160 x 160 possible combinations. Figure 9 shows the KS statistics plotted against Chi-squared for all 25600 points, with Chi-squared on a log scale because of the range of values. To be consistent with the previous figure, the red circles correspond to cases where both the Chi-squared and the KS statistic were significant and the yellow ones refer to cases where the Chi-squared was significant but the KS was not. The blue circles correspond to paired sets where neither statistic was significant, that is, the empty circles in the previous figure.

**Fig 9.**
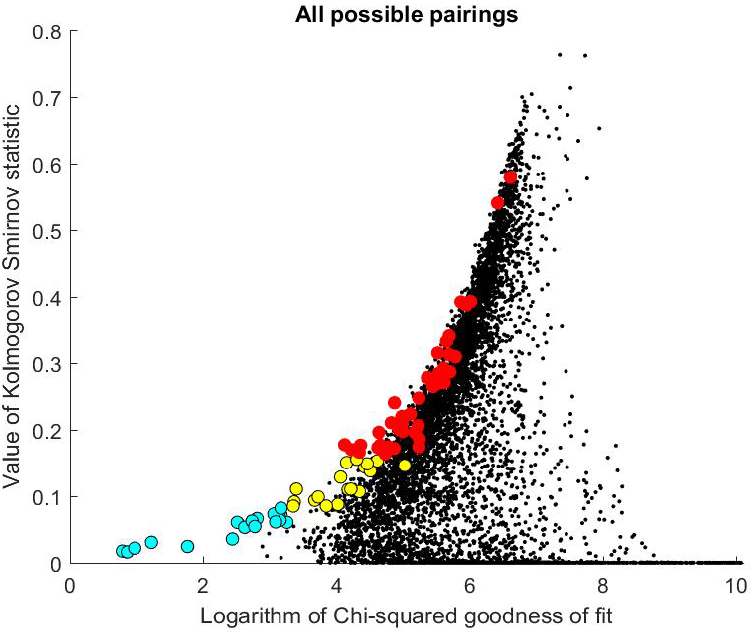
Kolmogorov-Smirnov statistic vs the chi-squared goodness of fit. In contrast to the previous figure, the two statistics were computed for every control simulation combined with every treatment simulation (i.e. 160 x 160). The chi-squared goodness of fit has been plotted on a log-scale.The aqua circles correspond to pairs of simulations where neither the chi-squared goodness of fit (Chi2) nor the Kolmogorov-Smirnov statistic (KS) are significant; the red circles, to cases where both are significant and the yellow circles, to cases where the chi-squared was significant but the KS was not.The paired simulations lie along the left-hand edge of the figure, and are similar to Markowitz’s efficient frontier in portfolio theory. [67]

Note how the circles corresponding to the paired simulations lie along the left-hand edge of the plot like a Markowitz efficient frontier [67]. The points along that frontier satisfy the condition that these have the lowest Chi-squared value for any given KS value. Or conversely they have the highest KS value for any Chi-squared value. The advantage of pairing the simulations is that it captures only the effect of the treatment and excludes the variability between control simulations.

### 6.4 Treatment effectively stops the epidemic

In the ideal case the treatment stops the epidemic in its tracks. This occurred in four sets of paired simulations; that is, in 2.5% of the cases.

## 7 Conclusion

In this paper we consider the problem of deciding whether or not to introduce a new treatment to prevent epidemics such as Zika, yellow fever, dengue etc. Decision-makers have to weigh up the potential advantages of the proposed treatment against its cost. *In silico* experiments can be used to test different strategies quickly and inexpensively.

Amongst the range of different simulation methods that are available now, agent-based simulations are emerging as a powerful tool in computational biology because they can capture fine scale heterogeneities and can reproduce the effects of interpersonal networks that can have important effects on the propagation of epidemics.

However given the inherent variability within the set of control simulations and within the set of simulations with treatment, it can be difficult to statistically prove their effectiveness. To overcome this problem, we have developed the concept of *paired simulations* - similar to paired t-tests. Paired simulations are generated using exactly the same random variables in the control simulation and one with a treatment.This approach is a powerful method for testing whether a given treatment is statistically better than the control, because it effectively filters out the differences within the control simulations and within the treatment simulations, leaving only the effect caused by the treatment.

This procedure has been applied to a case-study to see whether enclosing and air-conditioning the transport hub in Rio de Janeiro would have slowed down the propagation of Zika by eliminating the mosquito population there. Three types of statistics were compared for testing the hypothesis: Pearson’s chi-squared goodness of fit, the Kolmogorov-Smirnov statistic, and the Kullback-Leibler Divergence. The Kullback-Leibler Divergence performed poorly. This was to be expected because it is a constant multiple of the classical log likelihood ratio and Cressie and Read [29] had found that the latter performs well only when the alternative hypothesis has a dipped distribution (which is not our case). So the choice comes back to the Kolmogorov-Smirnov statistic or Pearson’s Chi-squared goodness of fit. To the best of our knowledge, no research has compared the power of these two statistics. Our tests on the sensitivity, precision and specificity of the two tests presented in Table 3 showed that the KS test although less sensitive, is much more precise and specific. **Consequently we recommend the Kolmogorov-Smirnov test**.

Although these new paired simulations were tested on the propagation of a vector-borne disease, they could be used to test the effectiveness of any treatment against epidemics and also in a wide range of other situations.

## Data Availability

This works does not include any data. Simulation Code is available on Github.

https://github.com/fccoelho/PairedSimulations

## Acknowledgments

- Health Secretariat of the City of Rio de Janeiro for providing the data;
- Dr Eduardo Mendes, EMAp Fundação Getulio Vargas, for his help
- Former civil engineer, C Leal, who had worked as an intern in designing the metro system in the Zona Sul, for his insights into the transport system;
- Participants of the Seminar on InfoDengue held at FGV on 19 October 2016, where a preliminary version of the work was presented, for the constructive comments.

## A Appendix: Supporting Information

### A.1 Summary of Zika Case Study

In October 2015, the Brazilian Ministry of Health (MoH) notified the World Health Organization of an unusual increase in the number of microcephaly cases among newborns in the state of Pernambuco, northeastern Brazil. On 28 November the MoH officially announced that there was a link between microcephaly and the Zika virus.

From mid-2015 to the end of January 2016, 47 cases of microcephaly were observed in the city of Rio de Janeiro, that were not due to other viral infections (syphilis, toxoplasmosis, herpes and cytomegalovirus). As these children were conceived from Dec 2014 to April 2015, Zika must have been rampant in the city from late 2014 onward, well before cases started to be recorded officially in October 2015.

#### A.1.1 Geographic spread within the city of Rio de Janeiro

Rio de Janeiro is divided into 160 areas called bairros which are regrouped into 33 administrative regions (RA for short). Their layout is given in Fig 10. The well-known ocean beaches (Copacabana and Ipanema) are in Zona Sul. Figure 11 shows the cumulative number of microcephaly cases due to Zika month by month from October 2015 until January 2016. Another reason for limiting the study to the end of January is that as people became aware that Zika caused microcephaly, women who were thinking of becoming pregnant started modifying their behavior to reduce the chances of catching Zika.

**Fig 10.**
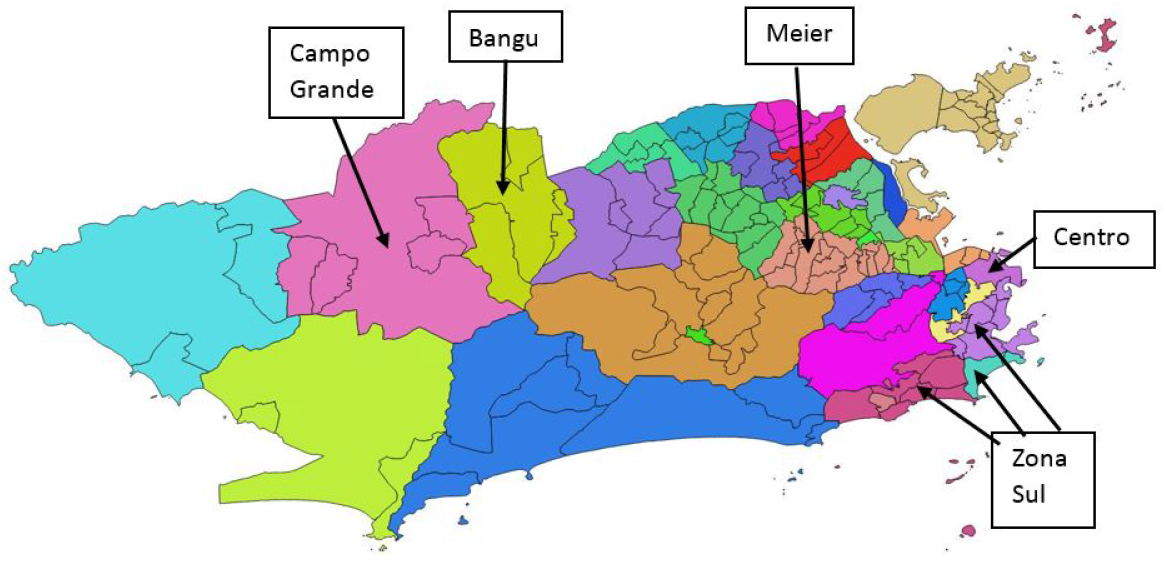
Location of the 33 administrative regions (RAs) in Rio de Janeiro. The five RAs that will be used for the simulation study are also shown.

**Fig 11.**
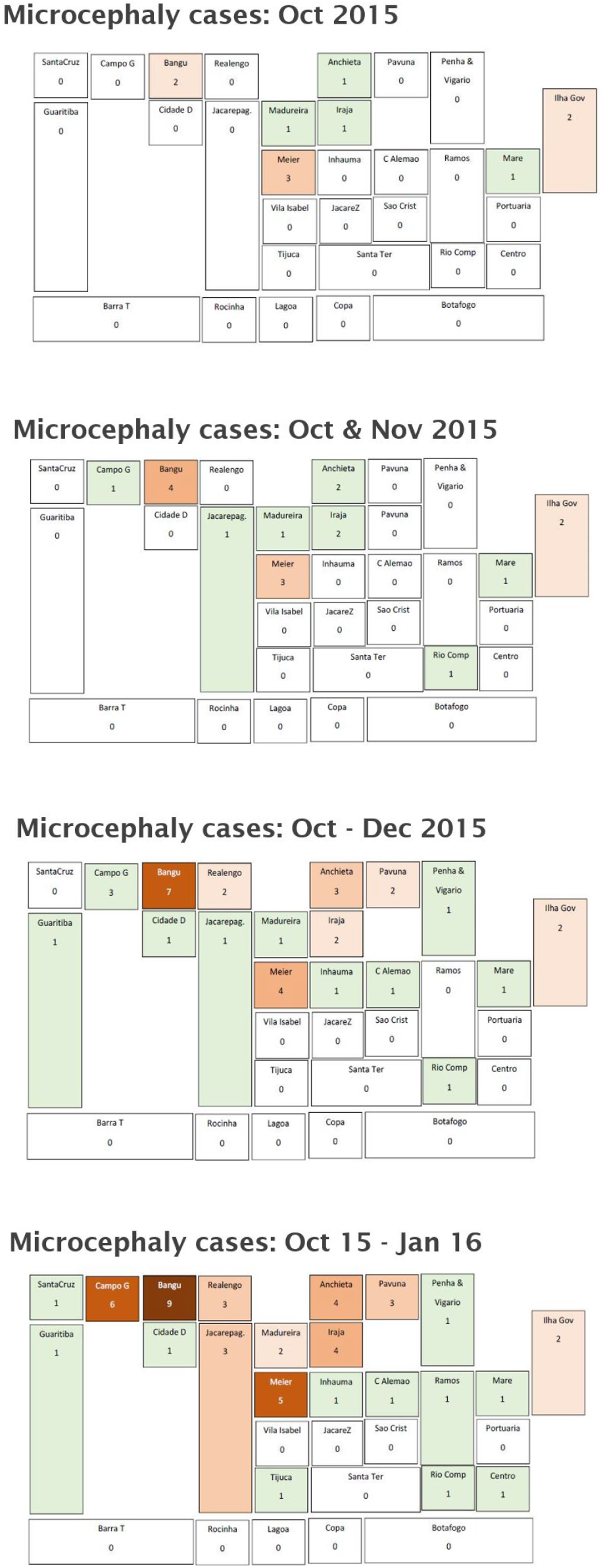
Microcephaly cases in Oct 2015 in Rio de Janeiro. The five RAs that will be used for the simulation study are also shown.

Cases were not evenly spread in proportion to the number of births; they were preferentially located in the northern suburbs, apparently following the public transport routes, with virtually no cases in favelas and none in the southern suburbs (Zona Sul). One key difference between the transport systems in the northern and southern suburbs is that the metro and rail system in the north is above ground whereas in the southern part the metro is underground with air-conditioning in carriages and forced ventilation on the platforms. The train system does not extend to Zona Sul.

#### A.1.2 Testing whether this spatial distribution could be random

Two separate hypotheses were tested to determine whether this spatial distribution could have occurred by chance. First we determined the probability of having zero cases in the Zona Sul out of the 47 microcephaly cases if they had been distributed randomly in proportion to the number of live births in the area. This hypothesis was rejected. Next after excluding Zona Sul, we computed the probability of 33 of the 47 cases occurring in the 9 RAs (Santa Cruz, Campo Grande, Bangu, Realengo, Anchieta, Pavuna, Madureira, Iraja and Meier) that lie along the above ground train and metro lines. Again, the hypothesis was rejected.

#### A.1.3 Testing whether socio-economic factors could explain the spatial distribution

The website of the city of Rio de Janeiro provides detailed information on the population and the living conditions in each RA, including a Human Development Index (HDI). In the 1990s, the UNDP developed a measure of human development that covered three aspects: longevity, education and economic development. Here we used the economic index which varies on a scale from 0 to 1 and measures the standard of living. First we computed the number of microcephaly cases per 1000 live births as a function of the HDI. The areas with the most microcephaly cases turned out to have an intermediate Human Development Index; areas with zero or 1 cases were either affluent areas in the Zona Sul or favelas which are much more disadvantaged. A linear regression model was fitted to test the hypothesis that the number of microcephaly cases per 1000 live births depends linearly on the municipal human development index, against the null hypothesis of a constant. This hypothesis was rejected: that is, the number of microcephaly cases per 1000 live births did not depend on the economic human development index.

#### A.1.4 On the metro and rail system in Rio de Janeiro

The public transport in Rio de Janeiro consists of train lines which are above ground and which take passengers from the northern suburbs into the city center (Fig 12) and metro lines which are above ground from the city center out north to Pavuna and below ground from the city center to Zona Sul in the southern suburbs (Fig 13), together with bus routes. Most of microcephaly cases occurred in RAs that lie along the train lines which are above ground, or along the metro lines in the northern part of the city which are also above ground. Surprisingly, areas in Zona Sul along the metro lines where the lines are underground have no cases.

**Fig 12.**
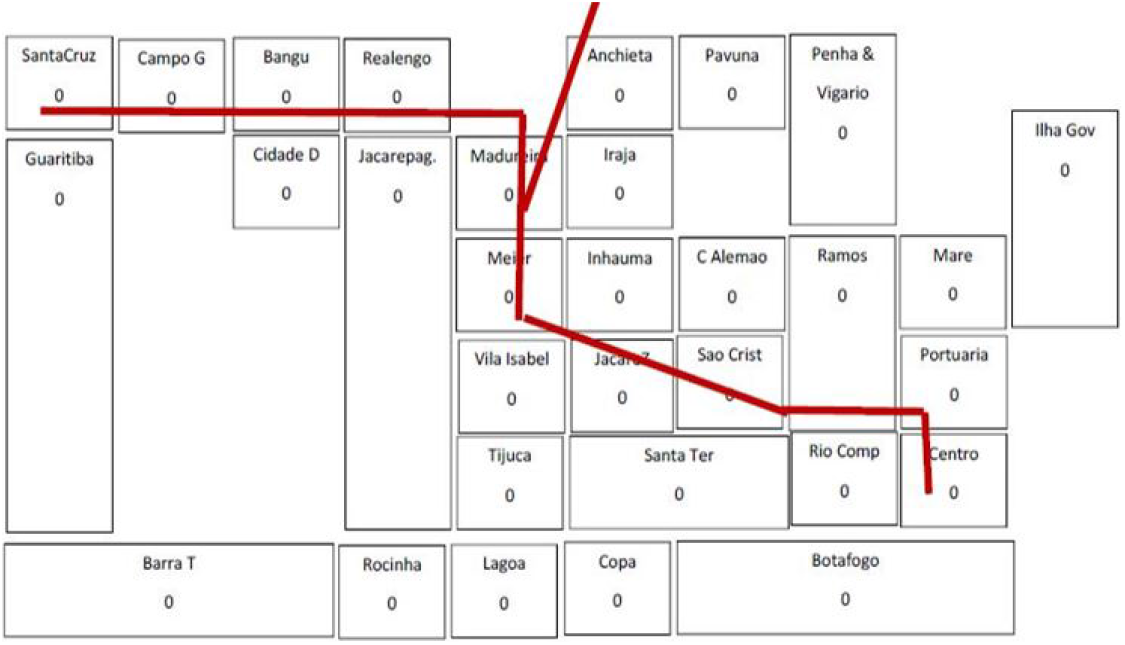
Train lines in Rio de Janeiro. Most cases of microcephaly were in areas along the train lines, which suggested that the transport system helped propagate the disease

**Fig 13.**
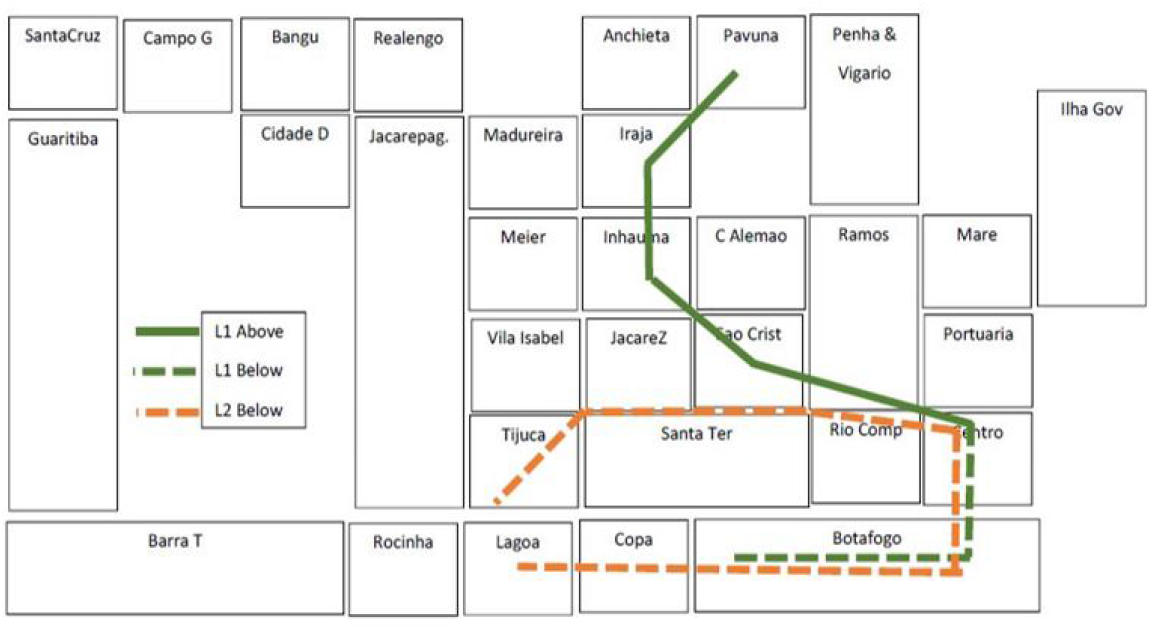
Metro lines in Rio de Janeiro. Cases also occured along the above ground part of the metro line, but surprisingly not along the underground part of Line 1 in Zona Sul

Anyone who travels on the metro in Rio notices how cold it is in the carriages and how windy it is on the platforms. We were curious to find out why it had been built this way. Was the aim to keep mosquitoes away or was this just a lucky consequence? The answer was provided by a former civil engineer who had worked on designing the stations soon after graduating. It had nothing to do with mosquito prevention. The fans and the air-conditioning in the carriages are for passengers’ comfort. Added to this, when trains go through tunnels, they push a wedge of air through in front of them, rather like a piston does, and large fans on the platforms increase the circulation of the air. A side effect is that it makes it difficult for mosquitoes to bite people.

### A.2 Simulating the impact of the transport system on the propagation of the Zika virus

It is widely recognized that human mobility is an important factor in spreading arboviruses such as dengue fever, chikungunya and zika. We wanted to test the hypothesis that air-conditioning in metro carriages and ventilation on the metro platforms in Zona Sul has made it more difficult for mosquitoes to bite people waiting on the platforms and hence led to a decrease in microcephaly cases in the southern suburbs. We used agent-based simulations to test this on a simplified (“toy”) example consisting of 5 RAs in Rio de Janeiro:

- three in the northern suburbs along the above ground train lines (Bangu, Campo Grande & Meier) which had many microcephaly cases,
- the city center with the main business area (Centro) and the main transport hub
- and the southern suburbs (Zona Sul).

Following Barmak et al. [12] and Barmak, Dorso & Otero [13] we divided each of these 5 RAs into two areas: the area around the train/metro station and the rest of the RA. This gave us 10 sub-zones each with its own mosquito population and its human population. or simplicity, we assumed the 10 mosquito populations are distinct and do not mix. According to the 2010 census there were only 41,00O people living in the Central area compared to 413,000 in Bangu, 542,000 in Campo grande, 398,000 in Meier and 569,000 in Zona Sul. So we ignored their presence in our model. The populations in the other four areas were split into two groups: those that go to the city center to work each day and those who remain in the home area. The latter go to work or to school there or they stay at home. People who stay in their residential area mix freely with each other but not with those from other areas. In contrast, people who go to work in the business area, mix with others there while doing business and while having lunch, and then again on the platform while waiting to go home.

#### A.2.1 Interactions between mosquitoes and people

We divided each day into 4 periods: early morning (when those who go to the city center wait on the platforms and get bitten by the station mosquitoes), midday (when people are in contact with either those in the city center, or with those in their home area), late afternoon when the travellers wait on the platforms at Centro and are bitten by that population of mosquitoes) and finally the evening when everyone is at home and gets bitten by the population of mosquitoes in that area.

Table 4 summarises the interactions between the 10 mosquito populations (M1 to M10) and the 8 populations of people (P1 to P6, P9 & P10). For example, the populations P1 + P3 + P5 + P9 wait for a train home late in the afternoon and can be bitten by mosquitoes from the M7 population. In the evening, populations P1 + P2 are at home in the Bangu area and can be bitten by mosquitoes from population M2.

#### A.2.2 Model parameters

We considered a SEIR model (susceptible, exposed, infectious and recovered) for people and a SEI model for mosquitoes. Mosquitoes do not recover; but they do die. As we only consider a period of several months, we ignored human mortality and people moving their residence from one area to another. When a susceptible mosquito bites an infectious person the virus is not always transmitted to it, and similarly when an infectious mosquito bites a susceptible person. These transmissions are controlled by two probabilities. The main parameters in the model listed in Table 5 were taken from Bastos et al (2016). When a mosquito dies, it is replaced by a new susceptible mosquito. We did not allow for vertical transmission between mosquitoes and their progeny. For simplicity, the incubation periods and the time to recovery were taken to be deterministic.

#### A.2.3 Simulation procedure

In our toy example we assumed that each mosquito population M1 to M10 consists of 10 mosquitoes and that there are 10 people in the each of the populations P1 to P6, P9 and P10. So there are 20 inhabitants in Bangu in the model: 10 who travel to the city center each work day and 10 who remaining in the Bangu area. The simulation was run three times for a total of 120 days, firstly with mosquito populations M1 to M10, secondly with no mosquitoes on the metro station Zona Sul, and thirdly with no mosquitoes at Centro.

Fig 14 shows the total number of cases in Zona Sul for the base case (red) and the case when there are no mosquitoes in the station area. Removing the mosquitoes from the station at Zona Sul (but leaving them in the Zona Sul living area) clearly reduces the number of cases throughout the city, but removing them from the transport hub at Centro is far more effective (Fig 15).

**Fig 14.**
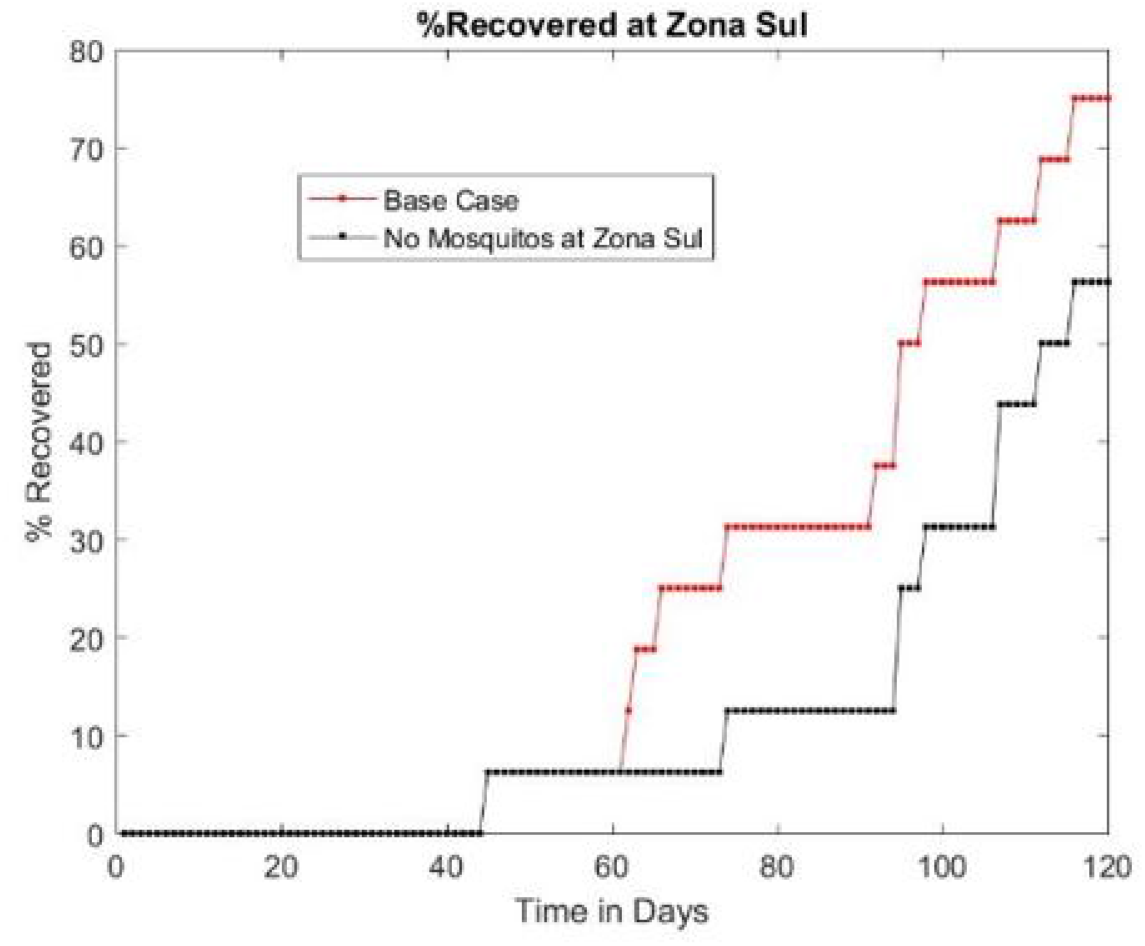
Total number of cases after 120 days in Zona Sul (out of a maximum of 40, in the base case (red), with no mosquitoes in Zona Sul (black)

**Fig 15.**
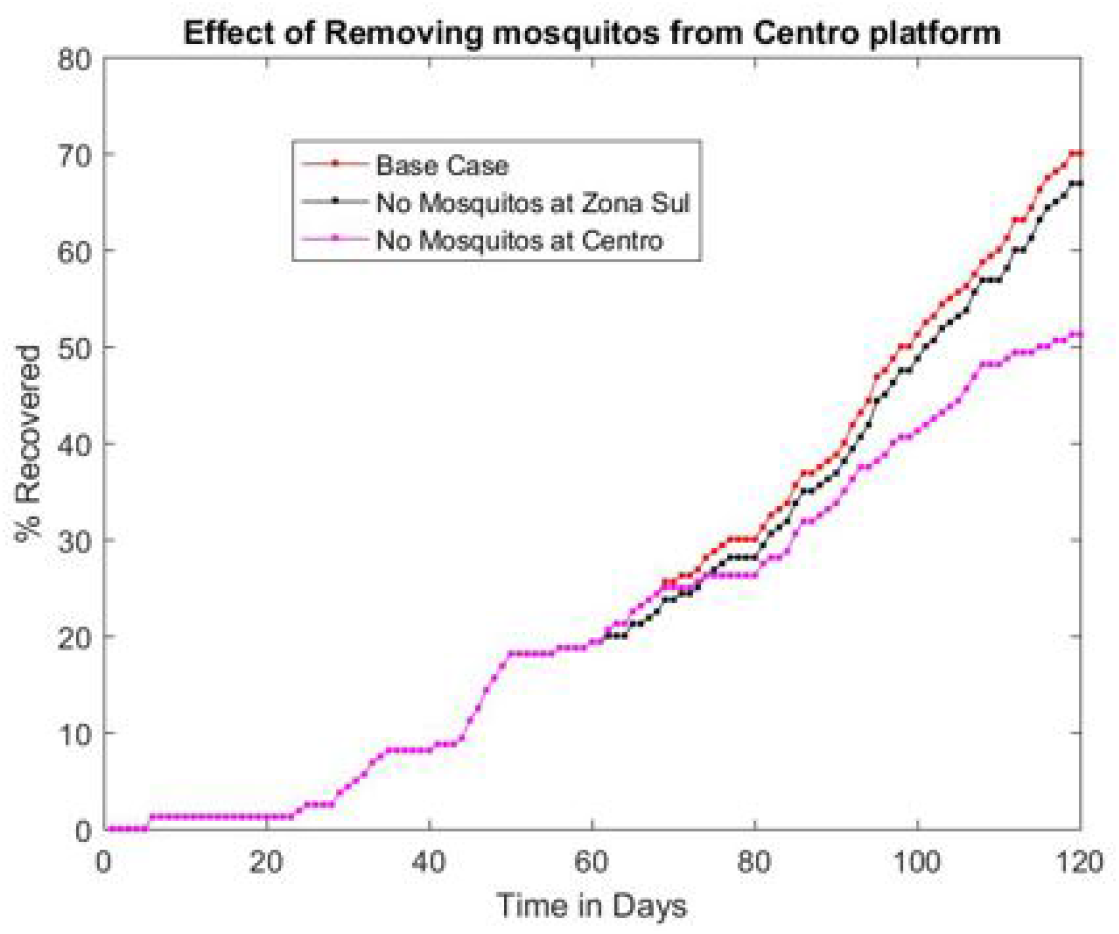
Results of three simulations. Total number of cases after 120 days (out of a maximum of 160, in the base case (red), with no mosquitoes in Zona Sul (black) and with no mosquitoes at Centro (mauve)

## Footnotes

1 Sexual transmission from men to women does occur but it is secondary compared to transmission by mosquitoes [3], [4].

2 Confirmed and probable cases, and those still under investigation by the Rio de Janeiro Health Secretariat.

3 http://nowak.ece.wisc.edu/ece830/ece830_lecture7.pdf

## Notes

### Competing Interest Statement

The authors have declared no competing interest.

### Funding Statement

No external funding is associated with this work.

## References

1. Rasmussen SA, Jamieson DJ, Honein MA, Petersen LR. Zika virus and birth defects—reviewing the evidence for causality. New England Journal of Medicine. 2016;374(20):1981–1987.

2. Cao-Lormeau VM, Blake A, Mons S, Lastére S, Roche C, Vanhomwegen J, et al. Guillain-Barré Syndrome outbreak associated with Zika virus infection in French Polynesia: a case-control study. The Lancet. 2016;387(10027):1531–1539.

3. Coelho FC, Durovni B, Saraceni V, Lemos C, Codeco CT, Camargo S, et al. Higher incidence of Zika in adult women than adult men in Rio de Janeiro suggests a significant contribution of sexual transmission from men to women. International Journal of Infectious Diseases. 2016;51:128–132.

4. D’Ortenzio E, Matheron S, de Lamballerie X, Hubert B, Piorkowski G, Maquart M, et al. Evidence of sexual transmission of Zika virus. New England Journal of Medicine. 2016;374(22):2195–2198.

5. Honório NA, Silva WdC, Leite PJ, Gonçalves JM, Lounibos LP, Lourenço-de Oliveira R. Dispersal of Aedes aegypti and Aedes albopictus (Diptera: Culicidae) in an urban endemic dengue area in the State of Rio de Janeiro, Brazil. Memórias do Instituto Oswaldo Cruz. 2003;98(2):191–198.

6. Trpis M, Hausermann W. Dispersal and other population parameters of Aedes aegypti in an African village and their possible significance in epidemiology of vector-borne diseases. The American journal of tropical medicine and hygiene. 1986;35(6):1263–1279.

7. Service M. Mosquito (Diptera: Culicidae) dispersal—the long and short of it. Journal of medical entomology. 1997;34(6):579–588.

8. Kraemer MU, Reiner RC, Brady OJ, Messina JP, Gilbert M, Pigott DM, et al. Past and future spread of the arbovirus vectors Aedes aegypti and Aedes albopictus. Nature microbiology. 2019; p. 1.

9. Stoddard ST, Morrison AC, Vazquez-Prokopec GM, Soldan VP, Kochel TJ, Kitron U, et al. The role of human movement in the transmission of vector-borne pathogens. PLoS neglected tropical diseases. 2009;3(7):e481.

10. Stoddard ST, Forshey BM, Morrison AC, Paz-Soldan VA, Vazquez-Prokopec GM, Astete H, et al. House-to-house human movement drives dengue virus transmission. Proceedings of the National Academy of Sciences. 2013;110(3):994–999.

11. Stone CM, Schwab SR, Fonseca DM, Fefferman NH. Contrasting the value of targeted versus area-wide mosquito control scenarios to limit arbovirus transmission with human mobility patterns based on different tropical urban population centers. PLoS neglected tropical diseases. 2019;13(7):e0007479.

12. Barmak DH, Dorso CO, Otero M, Solari HG. Dengue epidemics and human mobility. Phys Rev E. 2011;84:011901. doi:10.1103/PhysRevE.84.011901.

13. Barmak DH, Dorso CO, Otero M. Modelling dengue epidemic spreading with human mobility. Physica A: Statistical Mechanics and its Applications. 2016;447:129–140.

14. Hethcote HW. The mathematics of infectious diseases. SIAM review. 2000;42(4):599–653.

15. Eubank S, Guclu H, Kumar VA, Marathe MV, Srinivasan A, Toroczkai Z, et al. Modelling disease outbreaks in realistic urban social networks. Nature. 2004;429(6988):180.

16. Perez L, Dragicevic S. An agent-based approach for modeling dynamics of contagious disease spread. International journal of health geographics. 2009;8(1):50.

17. Maneerat S, Daudé E. A spatial agent-based simulation model of the dengue vector Aedes aegypti to explore its population dynamics in urban areas. Ecological Modelling. 2016;333:66–78.

18. Simoy M, Simoy M, Canziani G. The effect of temperature on the population dynamics of Aedes aegypti. Ecological modelling. 2015;314:100–110.

19. LaCon G, Morrison AC, Astete H, Stoddard ST, Paz-Soldan VA, Elder JP, et al. Shifting patterns of Aedes aegypti fine scale spatial clustering in Iquitos, Peru. PLoS neglected tropical diseases. 2014;8(8):e3038.

20. Isidoro C, Fachada N, Barata F, Rosa A. Agent-based model of Aedes aegypti population dynamics. In: Portuguese Conference on Artificial Intelligence. Springer; 2009. p. 53–64.

21. De Almeida SJ, Ferreira RPM, Eiras ÁE, Obermayr RP, Geier M. Multi-agent modeling and simulation of an Aedes aegypti mosquito population. Environmental modelling & software. 2010;25(12):1490–1507.

22. Dommar CJ, Lowe R, Robinson M, Rodó X. An agent-based model driven by tropical rainfall to understand the spatio-temporal heterogeneity of a chikungunya outbreak. Acta tropica. 2014;129:61–73.

23. Hunter E, Mac Namee B, Kelleher J. An open-data-driven agent-based model to simulate infectious disease outbreaks. PloS one. 2018;13(12):e0208775.

24. Macal CM, North MJ. Tutorial on agent-based modeling and simulation. In: Proceedings of the Winter Simulation Conference, 2005. IEEE; 2005. p. 14–pp.

25. Macal CM, North MJ. Tutorial on agent-based modeling and simulation part 2: how to model with agents. In: Proceedings of the 38th conference on Winter simulation. Winter Simulation Conference; 2006. p. 73–83.

26. Johnson NL, Leone FC. Statistics and experimental design: in engineering and the physical science. John Wiley; 1977.

27. McDonald JH. Handbook of biological statistics. vol. 2. sparky house publishing Baltimore, MD; 2009.

28. Whittle P. The outcome of a stochastic epidemic—a note on Bailey’s paper. Biometrika. 1955;42(1-2):116–122.

29. Cressie N, Read TR. Multinomial goodness-of-fit tests. Journal of the Royal Statistical Society: Series B (Methodological). 1984;46(3):440–464.

30. Armstrong M, Coelho FC, Bastos L, Saraceni V, Lemos C, Silva M, et al. How Public Transport affected the Propagation of Zika and Microcephaly within Rio de Janeiro early in 2015. BioRxiv. 2018; p. 345454.

31. Dorratoltaj N, Marathe A, Lewis BL, Swarup S, Eubank SG, Abbas KM. Epidemiological and economic impact of pandemic influenza in Chicago: Priorities for vaccine interventions. PLoS computational biology. 2017;13(6):e1005521.

32. Ferguson NM, Cummings DA, Fraser C, Cajka JC, Cooley PC, Burke DS. Strategies for mitigating an influenza pandemic. Nature. 2006;442(7101):448.

33. Barrett CL, Beckman RJ, Khan M, Anil Kumar V, Marathe MV, Stretz PE, et al. Generation and analysis of large synthetic social contact networks. In: Winter Simulation Conference. Winter Simulation Conference; 2009. p. 1003–1014.

34. Meng Y, Davies R, Hardy K, Hawkey P. An application of agent-based simulation to the management of hospital-acquired infection. Journal of Simulation. 2010;4(1):60–67.

35. Li Z, Swann JL, Keskinocak P. Value of inventory information in allocating a limited supply of influenza vaccine during a pandemic. PloS one. 2018;13(10):e0206293.

36. Dalgıç Ö O, Özaltın OY, Ciccotelli WA, Erenay FS. Deriving effective vaccine allocation strategies for pandemic influenza: Comparison of an agent-based simulation and a compartmental model. PloS one. 2017;12(2):e0172261.

37. Abbate JL, Murall CL, Richner H, Althaus CL. Potential impact of sexual transmission on Ebola virus epidemiology: Sierra Leone as a case study. PLoS neglected tropical diseases. 2016;10(5):e0004676.

38. Gerardin J, Ouédraogo AL, McCarthy KA, Eckhoff PA, Wenger EA. Characterization of the infectious reservoir of malaria with an agent-based model calibrated to age-stratified parasite densities and infectiousness. Malaria journal. 2015;14(1):231.

39. Pizzitutti F, Pan W, Barbieri A, Miranda JJ, Feingold B, Guedes GR, et al. A validated agent-based model to study the spatial and temporal heterogeneities of malaria incidence in the rainforest environment. Malaria journal. 2015;14(1):514.

40. Bomblies A. Agent-based modeling of malaria vectors: the importance of spatial simulation. Parasites & vectors. 2014;7(1):308.

41. Manore CA, Hickmann KS, Xu S, Wearing HJ, Hyman JM. Comparing dengue and chikungunya emergence and endemic transmission in A. aegypti and A. albopictus. Journal of theoretical biology. 2014;356:174–191.

42. Shi ZZ, Wu CH, Ben-Arieh D. Agent-based model: a surging tool to simulate infectious diseases in the immune system. Open Journal of Modelling and Simulation. 2014;2(01):12.

43. Voliotis M, Thomas P, Grima R, Bowsher CG. Stochastic simulation of biomolecular networks in dynamic environments. PLoS computational biology. 2016;12(6):e1004923.

44. Holmes AB, Kalvala S, Whitworth DE. Spatial simulations of myxobacterial development. PLoS computational biology. 2010;6(2):e1000686.

45. Wambaugh J, Shah I. Simulating microdosimetry in a virtual hepatic lobule. PLoS computational biology. 2010;6(4):e1000756.

46. Barrett C, Bisset K, Chandan S, Chen J, Chungbaek Y, Eubank S, et al. Planning and response in the aftermath of a large crisis: An agent-based informatics framework. In: Proceedings of the 2013 Winter Simulation Conference: Simulation: Making Decisions in a Complex World. IEEE Press; 2013. p. 1515–1526.

47. Lewis B, Swarup S, Bisset K, Eubank S, Marathe M, Barrett C. A simulation environment for the dynamic evaluation of disaster preparedness policies and interventions. Journal of public health management and practice: JPHMP. 2013;19(0 2):S42.

48. Niida A, Hasegawa T, Miyano S. Sensitivity analysis of agent-based simulation utilizing massively parallel computation and interactive data visualization. PloS one. 2019;14(3):e0210678.

49. Rizzi RL, Kaizer WL, Rizzi CB, Galante G, Coelho FC. Modeling Direct Transmission Diseases Using Parallel Bitstring Agent-Based Models. IEEE Transactions on Computational Social Systems. 2018;5(4):1109–1120.

50. Rizzi RL, Kaizer WL, Rizzi CB, Galante G, Coelho FC. A bitstring approach for implementing agent-based epidemiological models. Multiagent and Grid Systems. 2017;13(4):353–371.

51. Beckman RJ, Baggerly KA, McKay MD. Creating synthetic baseline populations. Transportation Research Part A: Policy and Practice. 1996;30(6):415–429.

52. Chowell G, Hyman JM, Eubank S, Castillo-Chavez C. Scaling laws for the movement of people between locations in a large city. Physical Review E. 2003;68(6):066102.

53. Salathe M, Bengtsson L, Bodnar TJ, Brewer DD, Brownstein JS, Buckee C, et al. Digital epidemiology. PLoS computational biology. 2012;8(7):e1002616.

54. Farooq B, Bierlaire M, Hurtubia R, Flötteröd G. Simulation based population synthesis. Transportation Research Part B: Methodological. 2013;58:243–263.

55. Bastos L, Villela DAM, Carvalho LM, Cruz OG, Gomes MFC, Durovni B, et al. Zika in Rio de Janeiro: Assessment of basic reproductive number and its comparison with dengue; p. 055475. doi:10.1101/055475.

56. Teukolsky SA, Flannery BP, Press W, Vetterling W. Numerical recipes in C. SMR. 1992;693(1):59–70.

57. Matsumoto M, Nishimura T. Mersenne twister: a 623-dimensionally equidistributed uniform pseudo-random number generator. ACM Transactions on Modeling and Computer Simulation (TOMACS). 1998;8(1):3–30.

58. Forsythe GE, Malcolm MA, Moler CB. Computer Methods for Mathematical Computations. Prentice Hall Professional Technical Reference; 1977.

59. Kullback S, Leibler RA. On information and sufficiency. The annals of mathematical statistics. 1951;22(1):79–86.

60. Kullback S. Statistics and Information theory; 1959.

61. Hoeffding W. Probability inequalities for sums of bounded random variables. In: The Collected Works of Wassily Hoeffding. Springer; 1994. p. 409–426.

62. Kolmogorov A. Sulla determinazione empirica di una lgge di distribuzione. Inst Ital Attuari, Giorn. 1933;4:83–91.

63. Kolmogorov AN. Foundations of probability. NY: Chelsea Publishing Co. 1933;.

64. Smirnov N. Table for estimating the goodness of fit of empirical distributions. The annals of mathematical statistics. 1948;19(2):279–281.

65. Massey Jr FJ. The Kolmogorov-Smirnov test for goodness of fit. Journal of the American statistical Association. 1951;46(253):68–78.

66. Fawcett T. An introduction to ROC analysis. Pattern recognition letters. 2006;27(8):861–874.

67. Markowitz H. Portfolio selection. The journal of finance. 1952;7(1):77–91.

